# Clinical Manifestations and Pregnancy Outcomes of Covid-19 in Indonesian Referral Hospital in Central Pandemic Area

**DOI:** 10.1101/2021.06.20.21259219

**Authors:** Muhammad Ilham Aldika Akbar, Khanisyah Erza Gumilar, Rino Andriya, Manggala Pasca Wardhana, Pungky Mulawardhana, Jimmy Yanuar Anas, Ernawati, Muhammad Ardian Cahya Laksana, Gustaaf Dekker

## Abstract

**Objectives:** The data on clinical manifestations and pregnancy outcomes of pregnant women with COVID-19 are limited, particularly in developing countries. The aim of this study is to analyze the clinical manifestations and pregnancy outcomes in COVID-19 maternal cases in a large referral hospital in Indonesia

**Methods:** The study used a prospective cohort design of all pregnant women with suspected COVID-19. Subjects were divided into COVID-19 and non COVID-19 group based on real-time polymerase chain reaction (RT-PCR) of SARS-CoV-2. The clinical characteristics, laboratory results, and pregnancy outcomes were then compared between both groups.

**Results:** From 141 suspected maternal cases, 62 COVID-19 cases were confirmed (43.9%), while 79 suspected cases were found to be negative (56.1%). The clinical manifestations and laboratory findings between the two groups were not significantly different (p>0.05). However, the maternal mortality directly caused by COVID-19 was significantly higher compared to the non-COVID-19 group (8.3 vs 1.3%; p=0.044; OR 6.91, 95% CI: 0.79-60.81).

**Conclusions:** The clinical manifestation and laboratory of suspected pregnant women with positive and negative RT-PCR COVID-19 result are similiar. However, within the Indonesian setting, COVID-19 strongly increases the risk of maternal death through both direct and indirect factors.

## Introduction

Since the first case of human Corona virus infection/ Corona disease (Covid-19) was announced in Wuhan (Dec 31st 2019), the virus has already infected more than 30 million people and lead to almost million deaths on Sept 30th 2020 (WHO Covid-19 Dashboard). WHO declared Covid-19 a pandemic disease in March 2020. Indonesia, as the third most populated country in Asia, has become one of the major COVID-19 hot spots in the Southeast Asia region. Data from Indonesia government (September 21, 2020) showed that the total number of COVID-19 cases are >245,000, with 9,553 total deaths and 177,000 recovered cases; although the accuracy of these national data is not certain. Estimation of the COVID-19 case fatality rate (CFR) in Indonesia is 3.9%. Indonesian COVID-19 cases are increasing sharply with the average number of confirmed new cases approximately 4.000-5.000/day since the first confirmed case on March 2, 2020.

Pregnant women with COVID-19 provided difficult challenges to the maternal health services in the East Java province, particularly in Surabaya. Most smaller and regional hospitals were not prepared to managed COVID -19 cases, mainly due to lack of facilities and infrastructure, availability of isolation rooms and personal protective equipment (PPE) [1]. At the beginning of pandemic, there were only four referral hospitals in Surabaya able to handle maternal COVID-19 cases. One of them is the Airlangga University Academic Hospital (RS UNAIR). RS UNAIR is the secondary hospital-level which has been appointed as one of the biggest referral COVID-19 hospitals in Surabaya. We handled the first cases of pregnancy with COVID-19 in mid-April 2020; the start of this cohort study.

To date, the effect of COVID-19 in pregnancy is still uncertain. Data on pregnant women with COVID-19 is limited and mostly based on a few cases. Wuhan’s researchers published one of the first reports about COVID-19 in pregnancy. This retrospective study reported was based on the medical records of 9 confirmed cases of COVID-19 pregnancy [2]. The study showed similar clinical characteristics of COVID-19 between pregnant and non-pregnant women with no evidence of intrauterine vertical transmission. Since then many papers have been published on COVID-19 during pregnancy; however, most of these are based on retrospective medical record reviews or case reports, most papers found results similar to the initial Wuhan series. Various systematic reviews published on larger sample sizes; 108 cases [3], 92 cases [4], 90 cases [5], and 37 cases [6] reported reassuring findings in term of clinical outcomes.

Based on the data above, this study aims to analyze the clinical manifestations and pregnancy outcomes (maternal-fetal) in a prospective cohort of COVID-19cases, in Airlangga University Academic Hospital (RS UNAIR).

## Material & Methods

This is a prospective cohort study of pregnant women with suspected COVID-19 cases in Universitas Airlangga Hospital (Surabaya, Indonesia) from April – August 2020. This study has been approved by Ethical Committee of Universitas Airlangga Hospital (Ref. N0:110/KEP/2021). Informed consent was obtained from all individual participants included before the study began. The subjects were recruited based on our screening methods, which is the presence of one of the clinical sign-symptoms of COVID 19 (fever, cough, dyspnea, odinophagia, myalgia, or nausea vomiting), or the positive rapid antibody COVID-19 test, or chest x-ray findings, or an abnormal complete blood count (CBC) especially the neutrophil/lymphocyte ratio (NLR) (>5.8) (7). The study involved all pregnant women with suspected of COVID-19 giving birth in UNAIR Hospital during April 15th the end of August 2020. All the suspected COVID-19 cases were divided based on the *Real Time Polymerase Chain Reaction* (RT-PCR) result as COVID versus non-COVID group (Figure 1). The RT-PCR sample was taken from nasopharyngeal and oropharyngeal swab. Patients were followed until delivery and the pregnancy outcomes data were collected. The primary outcomes of the study were clinical manifestations and pregnancy outcomes of COVID-19 in pregnancy. Pregnancy outcomes were divided into maternal outcomes (maternal death, gestational age at delivery, hospital length of stay, pregnancy complications, mode of delivery) and fetal outcomes (preterm delivery rates, birth weight,length, and 1 and 5 minute Apgar scores).

**Figure 1.**
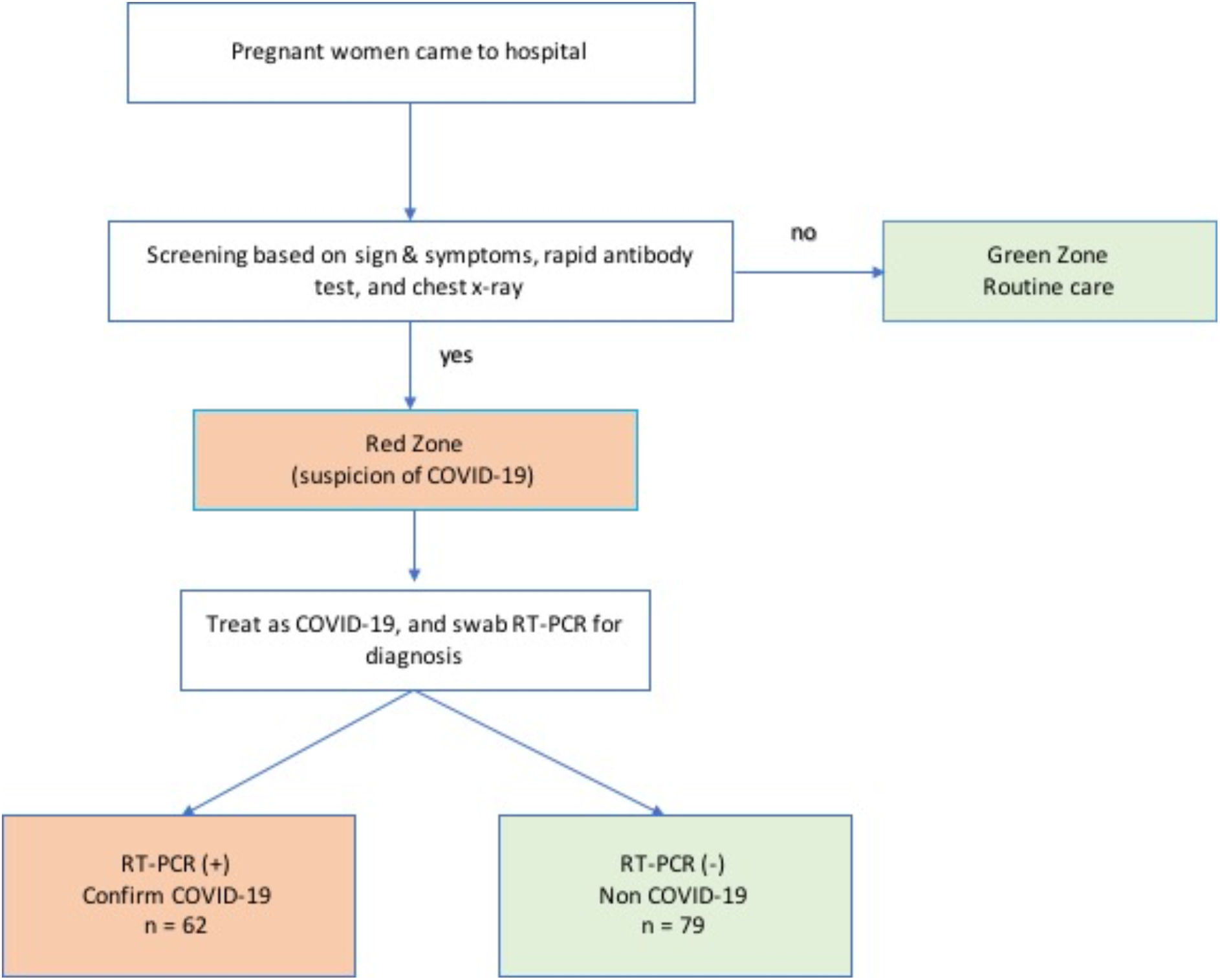
Algorithm of Patients Recruitment

The data was analysed using IBM^@^ SPSS^©^ Statistics ver 25. Descriptive statistics was used first to determine the distribution normality of the numerical variables. The numerical variables with normal distribution (hemogloblin, leucocytes, thrombocyte, and fetal birth weight) were analysed using independent t-test value. Other abnormal numerical variables distributed were applied to the Mann Whitney test. A Chi-Square test was used to analyse categorical variables. Furthermore, the categorical variables that do not fulfil the Chi-square test requirement were analysed by Fischer’s exact test (including contact status, myalgia, nausea-vomiting, and maternal outcomes). Significance was defined as p < 0.05.

## Results

We recruited a total 141 suspected COVID-19 cases in pregnancy during the study period; 62 cases (43.9%) were confirmed COVID-19, 79 were negative (56.1%).

### Maternal Background

Regarding baseline demographics no differences were identified between 2 groups (maternal ages, parity, antenatal care frequency, patients entry, referral origin, antenatal care provider, and comorbidities) (*p* > 0.05) (table 1). The only difference was that the COVID-19 group had a higher history of contact with sick patients compared to non COVID-19 group (6.5% vs 0). All contact sources were family members diagnosed with a confirmed COVID-19. Most patients were referred from other hospitals/clinics within the city. No patients had a travel history outside the city/country within 2 weeks before onset of the symptoms.

**Table 1.** Maternal Background.

|  | Non Covid | Covid | *p* value |
| --- | --- | --- | --- |
|  | n=79 | n=62 |  |
| Maternal Ages<sup>a</sup><br>(years old) | 28 (7) | 28.5 (8) | 0.118 |
| Parity<sup>a</sup> | 1 (1) | 1 (2) | 0.197 |
| ANC Frequency<sup>a</sup> | 8 (4) | 7.5 (3) | 0.412 |
| Patients Origin |  |  |  |
| Self Appointment | 24 (30.4%) | 17 (27.4%) | 0.825 |
| Referral | 21 (26.6%) | 18 (29%) |  |
| Free Referral | 28 (35.4%) | 23 (37.1%) |  |
| Outpatient Clinics | 5 (6.3%) | 2 (3.2%) |  |
| Other Department | 1 (1.3%) | 2 (3.2%) |  |
| ANC Provider |  |  |  |
| No ANC | 0 | 1 (1.6%) | 0.206 |
| Primary Health Care<br>Services | 2 (2.5%) | 4 (6.5%) |  |
| Hospital | 9 (11.4%) | 13 (21%) |  |
| Midwifery clinic | 5 (6.3%) | 2 (3.2%) |  |
| Mixed | 63 (79.7%) | 42 (67.7%) |  |
| Contact Status |  |  |  |
| No | 79 (100%) | 58 (93.5%) | 0.035* |
| Yes | 0 | 4 (6.5%) |  |

Note: \*indicate significant difference with $p$ value < 0.05. <sup>a</sup>indicate abnormal data distribution, present with a median (interquartile range). Abbreviation: ANC: Antenatal Care.

### Clinical Manifestation

The clinical manifestations and laboratory results between both groups were not statistically different (*p*>0.05) (Table 2). Only 24.2% pregnant women in the COVID-19 group had typical signs and symptoms, mostly in form of cough, fever, and dyspnea. We did not find anosmia and dysgeusia in all cases, which are considered as common signs of COVID-19. Complete blood counts (hemoglobin, leucocyte, thrombocyte, neutrophil, lymphocyte, and NLR) mean or median value were not significantly different between these two groups.

**Table 2.** Clinical Manifestation.

|  | Non Covid<br>n = 79 | Covid<br>n = 62 | *P* value |
| --- | --- | --- | --- |
| Sign & Symptoms | 19 (24.1%) | 15 (24.2%) | 0.984 |
| Cough | 16 (20.3%) | 12 (19.4%) | 0.894 |
| Fever | 5 (6.3%) | 9 (14.5%) | 0.107 |
| Dyspneu | 6 (7.6%) | 8 (12.9%) | 0.295 |
| Odinophagia | 4 (5.1%) | 3 (4.8%) | 0.951 |
| Myalgia | 0 | 1 (1.6%) | 0.440 |
| Nausea Vomitting | 2 (2.5%) | 4 (6.5%) | 0.405 |
| Rapid antibody test | 50 (64.9%) | 40 (72.7%) | 0.343 |
| Hemoglobin<sup>a</sup> (g/dL) | 11.34 ± 1.48 | 10.83 ± 1.80 | 0.072 |
| Leucocyte<sup>a</sup> (10<sup>3</sup>/uL) | 11270 ± 3500 | 10552 ± 5002 | 0.327 |
| Trombocyte<sup>a</sup> (10<sup>3</sup>/uL) | 279835 ± 83053 | 289306 ± 80894 | 0.504 |
| Neutrophil<sup>b</sup> | 76.1 (9.8) | 76.6 (11.75) | 0.217 |
| Lymphocyte<sup>b</sup> | 15.5 (7.15) | 15.8 (9.33) | 0.309 |
| NLR<sup>b</sup> | 4.88 (2.55) | 4.94 (4.15) | 0.226 |

Note: <sup>a</sup> indicate data with normal distribution, present with mean ± SD. <sup>b</sup> indicate data with abnormal distribution, present with median (interquartile range). Abbreviation: NLR: Netrophil/Lymphocyte Ratio.

### Pregnancy Outcomes

Maternal deaths by all causes in the COVID-19 group were significantly higher compared with the non COVID-19 group (9.8 vs 1.3%; p=0.044; OR 8.29, 95% CI: 0.97-70.84). The Odd Ratio (OR) of maternal deaths caused by ARDS of COVID-19 only were also significantly higher compared to non COVID-19 groups (8.3 vs 1.3 %; OR 6.91, 95% CI: 0.79-60.81). The length of stay in COVID-19 group was also higher compared to non COVID-19 (3 vs 2 days) patients. Majority of cases in both groups were delivered by cesarean section (63.3% and 71.7%), and about 5% was treated conservatively due to the preterm gestation. The three preterm cases of COVID-19 were treated conservatively for threatened preterm labour. The COVID-19 group tended to have a higher rate of pregnancy complications (such as hypertension in pregnancy, arrest of labor, misscariages, threatened preterm labor, post date pregnancy, oligohydramnions, and premature rupture of the membrane [PROM]) compared to non COVID-19 women (*p*=0.130). The prevalence of specific pregnancy complications in non COVID-19 group vs COVID-19 group are: hypertension in pregnancy (17.7% vs 16.1%), arrest of labour (6.33% vs 4.84%), and PROM (17.72% vs 11.29%), In addition, perinatal outcomes were not difference between both groups in term of birth weight, length, and Apgar scores. The rate of preterm delivery <37 weeks tended to be higher in COVID-19 pregnant women compared to non COVID (12.06% vs 6.49%) (table 3).

**Table 3.**
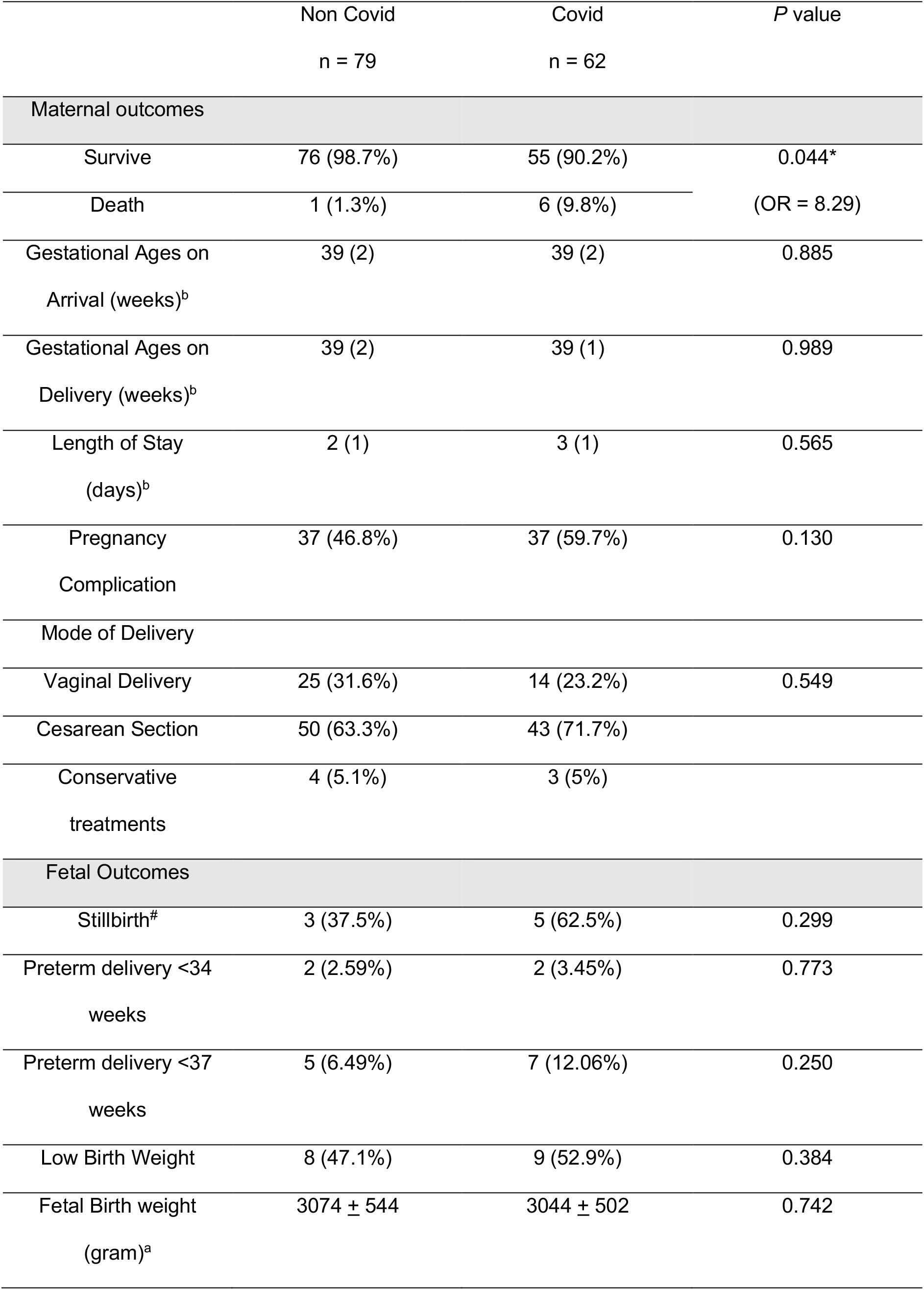
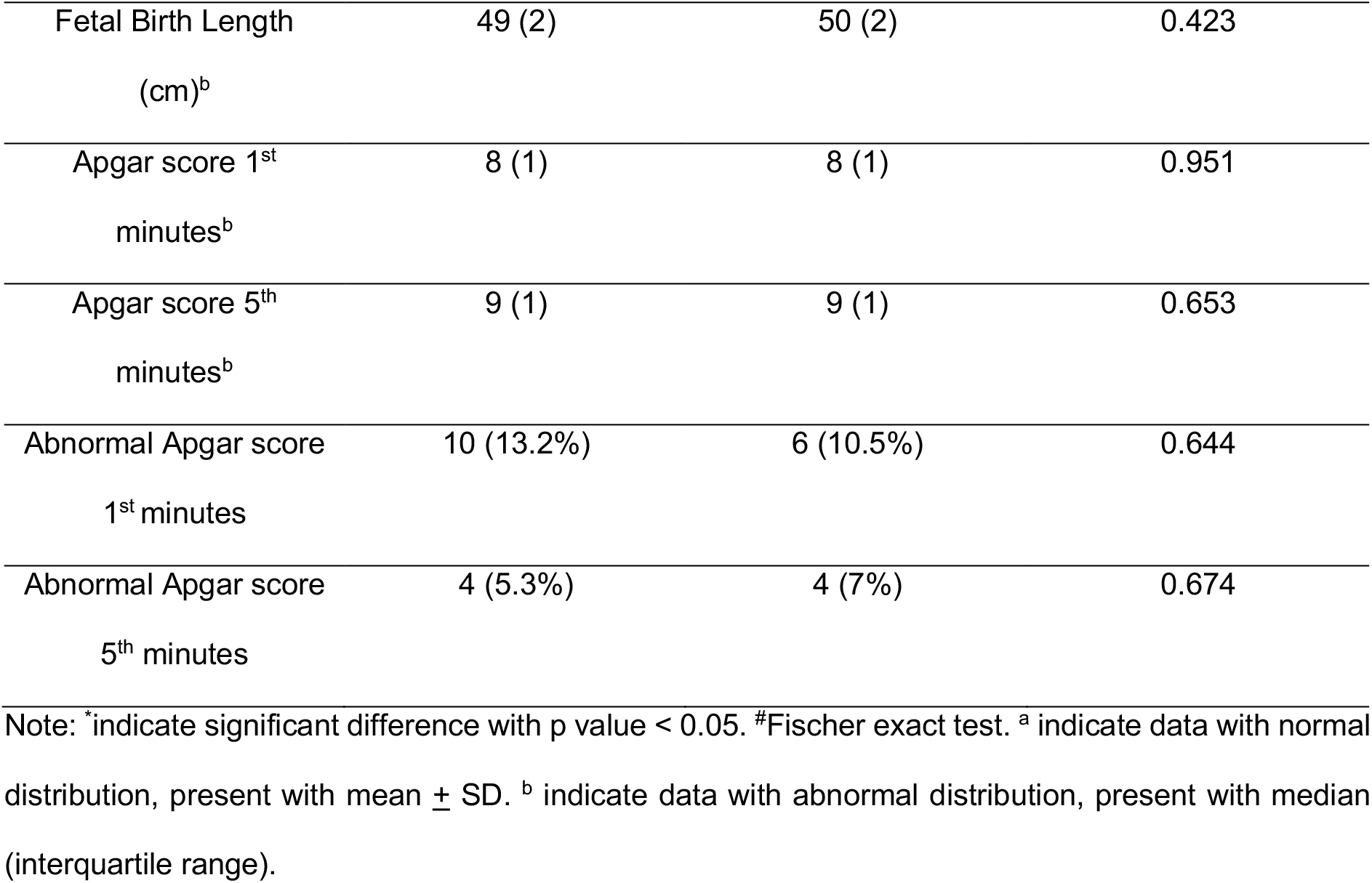
Maternal and Perinatal Outcomes.

## Discussions

The data of this prospective cohort of COVID 19 pregnant patients presenting with suspected COVID-19 in a major referral hospital in East Java demonstrated a significant higher risk of maternal death in COVID-19 positive cases. Interestingly only 24.2% of patient with confirmed COVID-19 had signs and symptoms during admission. In other words, the majority of COVID-19 cases on pregnancy were asymptomatic (75.8%), but were identified as suspected COVID-19 on the basis of antibody based screening test. This is consistent with the study findings in New York, which found 29 from 33 (87.9%) pregnant women with confirmed COVID-19 to be asymptomatic [8]. Based on these findings, COVID-19 universal screening for pregnant women in labour should be performed routinely in high local transmission disease areas such as Surabaya [8]. This is in line with the official national recommendation from POGI (Indonesia Obstetrics & Gynaecology Association) and also RCOG (The Royal College of Obstetricians and Gynaecology), to perform a universal screening for all delivering women [9]. COVID-19 status is important to determine a place of care, labour/post-partum/newborn management, the use of personal protective equipment (PPE), and the availability of isolation room. This policy is different from others guidelines such as FIGO, ISUOG, ISIDOG, RCPI recommending screening and diagnosis to be performed selectively in high risk cases, although these professional groups do acknowledge that the threshold of suspicion indicating of a lab based examination should be lowered [10–13]. The clinical sign and symptoms from this study include cough (19.4%), fever (14.5%), dyspnea (12.9%), nausea vomiting (6.5%), odynophagia (4.8%), and myalgia (1.6%). This finding is in line with many previous study [14–16].

The significantly higher risk of maternal death (>8x) in this prospective cohort is in contrast with the findings from the systematic review by Zaighman et al, and Smith V et al, which found no maternal death among total 200 pregnant women with COVID-19 [3,4]. Out of the 6 maternal death in this cohort, 5 were directly related to COVID-19, namely respiratory failure. Three cases involved term pregnancies, two were preterm, and the last one was in the 2^nd^ trimester (17 week’s gestation). One case was complicated with severe preeclampsia requiring cesarean hysterectomy because of severe haemorrhage caused by uterine atony uteri. Following the caesarean hysterectomy, the patient was treated in the ICU with ongoing mechanical ventilation for 10 days before her condition sadly deteriorated. The cause of death in this case was respiratory failure caused by COVID-19, complicated by severe preeclampsia and severe haemorrhage during labour. One maternal death was not caused directly by COVID-19: a term pregnant women was referred to our hospital because of COVID-19. The delivery was performed in private midwifery clinic, unfortunately fetal anencephaly was diagnosed and labour was complicated by a shoulder dystocia. This patient never had any antenatal care at all during pregnancy, and the COVID-19 test showed a positive result. The referral to our hospital was difficult because many hospitals capable of managing COVID-19 cases were full at that time. After one hour delay, the patient finally came to our hospital with the fetal head protruding, and already deceased caused by post partum haemmorhhage. This incident indicates that COVID-19 can also indirectly cause maternal death by delaying the referral or management of emergency obstetrics cases. In the non COVID-19 group one maternal death occurred due to multiple organ failure caused by an adverse reponse to the tuberculosis treatment she received.

The rate of maternal complication in COVID-19 group tended to be higher compared to non COVID-19 group (not statistically significant; 59.7% vs 46.8%). The prevalence of hypertension in pregnancy in COVID-19 group was not different with non COVID-19 group (16.1% vs 17.7%). Narang K et al, hypothesized that COVID-19 would enhance the risk of preeclampsia related to endothelial dysfunction and coagulation disorders induced by SARS-CoV 2 [17]. However in this cohort we did not find an increased preeclampsia risk.

The high caesarean section rate in the overall cohort is explained by the fact that particularly at the beginning of the pandemic, the view point prevailed that cesarean section had a lower risk of transmission to the medical staff. In addition the limited availability of PPE in many hospitals caused most hospital to choose cesarean section as a primary method of delivery related to the ability to conserve the PPE used [1]. Our national obstetrics gynecology association (POGI) also publish a first official recommendation in March 2020, to deliver a COVID-19 cases by cesarean section [18]. Cesarean section rate in our study is lower than the systematic review with 92 confirm COVID-19 pregnant women (80%) [4], and 108 cases (92%) [3]. So far, there is still lack of clear evidence which delivery methods is better in COVID-19 cases, in term of maternal and neonatal outcomes [19]. In a recent development, many international guidelines recommend that method and timing of delivery should be individualized, depending on the clinical status of the patients, gestational age, and the maternal and fetal condition [11,13,20]. Since August 2020, POGI publish a revision of the guideline and recommend that method of delivery should be decided based on the medical and obstetric indication [9]. What remains a concern about the delivery methods is the question if there is any difference for transmission of COVID-19 to the medical staff in cesarean section vs vaginal delivery. In a vaginal birth, mothers have to push during the second stage, which is an aerosol producing procedure that increase the number of droplets in the air, and eventually lead to higher risk of transmission to medical staff or birth attendant. This process does not occur in cesarean sections. To date, there is still no evidence about this matter, so future studies need to address this issue.

Preterm delivery < 37 weeks tend to be higher in COVID-19 group compared to non COVID-19, although not different statistically. Our preterm delivery < 37 weeks rate is much lower compare to other systematic review (12.06% vs 46.15%) [4]. The prevalence of preterm delivery < 34 weeks in COVID-19 group is 2.59%, which is not found in that systematic review [4]. In a multicenter cohort study (not yet published) by the Spanish Obstetrics Emergency Group, it is report that COVID-19 increase the risk of preterm delivery 2 fold (aOR 2.12, 95% CI: 1.32-3.36; p=0.002) [21]. The increasing preterm delivery risk is associated with cytokine storm in COVID-19, which induces an uncontrolled inflammatory response. Many inflammatory cytokines known to be increased in many cases of preterm birth are also increased in COVID-19, including IL-6, IL-1*β*, TNF-*α*, G-CSF, and macrophage [21]. Compared with the SARS pandemic (2003), our study reveal that the preterm delivery rate is lower. In one of the SARS series involving 12 pregnant in Hongkong, the preterm delivery rate was 33.3% [22]. Besides being caused by coronavirus, 2 from 7 preterm birth in this Indonesian series were iatrogenic because of severe preeclampsia.

The stillbirth rate was non-significantly higher in COVID-19 patients compared with the non COVID-19 group (62.5% vs 37.5%). There were five stillbirths in COVID-19 group, all related to maternal death. Of the five stillbirth cases in COVID-19 group, two babies were born preterm and with low birth weight. The number of stillbirths in our study in the COVID-19 group was much higher compared to any previous report. In a systematic review of 108 pregnancy with COVID-19, there were only 2 (1.85%) stillbirths [3]. In the other systematic review with 92 pregnant women with COVID-19, the perinatal mortality rate was 3.92% (2/37). All of this perinatal mortality was occurred in severe COVID-19 cases, which end in maternal death [4].

COVID-19 has been shown as a strong risk factor of maternal death in this study and this may contradict most available reports. Majority of maternal death is caused by the direct effect of COVID-19 respiratory failure), but maternal death may also be caused indirectly by delaying the referral and adequate management in hospital. Intensive and early management is required to prevent maternal death, by performing universal COVID-19 screening in all pregnant and delivering women.

## Data Availability

The data of this study are available from Universitas Airlangga Hospital, but restrictions apply to the availability of these data and so are not publicly available.

## Acknowledgements

We thank our Head and Secretary of Department, Brahmana Askandar Tjjokroprawiro and Ashon Saadi for their support in this research. We thank to our Hospital director and his staff, Prof. Nasronudin for their support in this study. We thank Erni Rosita Dewi for providing assistance in language correction. We thank all the research team for their remarkable effort in collecting data for this study. And we thank to all the patients who participate in this study.

## Ethical Approval

This study has been approved by Ethical Committee of Universitas Airlangga Hospital (Ref. N0:110/KEP/2021).

## Patient Consent

All study participants has given written consent to the inclusion of material pertaining to themselves, and authors have fully anonymized all participants.

## Conflict of Interest

Authors declare no conflict of interest in this study. This research received no spesific grant from any funding agency in the public, commercial, or not-for-profit sectors.

## Notes

### Competing Interest Statement

The authors have declared no competing interest.

### Funding Statement

There are no funding received for this study

